# Operant Conditioning of Rectus Femoris H-Reflexes: A Proof-of-Concept with Implications for Post-Stroke Quadriceps Hyperreflexia

**DOI:** 10.1101/2022.07.20.22277649

**Authors:** Kyoungsoon Kim, Tunc Akbas, Robert Lee, Kathleen Manella, James Sulzer

## Abstract

Hyperreflexia is common after neurological injury such as stroke, yet clinical interventions have had mixed success. Our previous research has shown that hyperreflexia of the rectus femoris (RF) during pre-swing is closely associated with reduced swing phase knee flexion in those with post-stroke Stiff-Knee gait (SKG). Thus, reduction of RF hyperreflexia may improve walking function in those with SKG after stroke. A non-pharmacological and non-surgical procedure for reducing hyperreflexia has emerged based on operant conditioning of spinal reflexes elicited using electrical stimulation of the peripheral nerve, known as an H-reflex. Operant H-reflex conditioning training of the soleus has improved clinical gait function in those with spinal cord injury. It is currently unknown whether operant conditioning can be applied to the RF or those after stroke. This feasibility study trained 7 participants (5 neurologically intact, 2 post-stroke) over a period of 3 months, up to 3 times per week, to down-condition the RF H-reflex using visual feedback. We found an overall decrease in average RF H-reflex amplitude among 7 participants (44% drop, p<0.001, paired t-test), of which the post-stroke individuals contributed (49% drop). We observed a generalized training effect across quadriceps muscles. Post-stroke individuals exhibited improvements in peak knee-flexion velocity, reflex excitability during walking, and clinical measures of spasticity. These aforementioned neuromuscular outcomes provide promising initial results that operant RF H-reflex conditioning is feasible both in healthy and post-stroke individuals. This procedure could provide a targeted alternative to spasticity management and avoid the drawbacks of drugs and surgery.

## Introduction

Stiff-Knee gait (SKG) after stroke is a common disability often defined by decreased knee flexion angle during the swing phase of walking (Perry and Burnfield 2010). Those with SKG suffer from joint pain, energy inefficiency and increased risk of falls (Burpee and Lewek 2015). The causes of post-stroke SKG are unclear, although it has been suggested that quadriceps spasticity contributes to SKG (Perry 1992). Our recent work demonstrates this claim, showing an association between hyperreflexia, a component of spasticity, in the rectus femoris (RF) and reduced knee flexion angle in those with post-stroke SKG (Akbas et al. 2020). Hyperreflexia is defined as overactive or overresponsive reflex activity (Lance 1980; Young and Wiegner 1987), possibly resulting from disinhibition of brainstem pathways such as the reticulospinal tract (Li and Francisco 2015; Li et al. 2018). Thus, a reduction of RF hyperreflexia may help restore healthy gait through restoration of spinal inhibitory pathways in those with post-stroke SKG.

Various pharmacological and surgical interventions have been used to treat quadriceps spasticity. Botulinum neurotoxin (BoNT) injection blocks neurotransmitter release at the neuromuscular junction, resulting in increased range of motion (ROM) in the knee joint angle (Robertson et al. 2009; Roche et al. 2015). BoNT injections alleviate hyperreflexia (Stoquart et al. 2008; Robertson et al. 2009; Roche et al. 2015), but also weaken the quadriceps throughout the gait cycle, resulting in limited clinical benefit in those with SKG (Stoquart et al. 2008; Wissel et al.), Surgical interventions such as tendon transfer surgery is also possible, but there is limited evidence for clinical effectiveness (Platz 2021). Baclofen, a gamma-Aminobutyric acid (GABA) agonist, is the most common oral treatment for spasticity, however its effectiveness is questionable and results in side effects such as sedation (Dehnadi Moghadam et al. 2021; Navarrete-Opazo et al. 2016). Baclofen is more effective when administered intrathecally in a targeted manner (Kofler et al. 2009; Dvorak et al. 2011), however it may negatively affect compensatory motor systems and requires a surgical implant with regular maintenance. While pharmacological and surgical interventions for hyperreflexia exist, they all have significant drawbacks.

A possible new non-pharmacological, non-surgical procedure could help target hyperreflexia using operant conditioning. Operant conditioning is a type of associative learning process through which the strength of a behavior is modified by reinforcement or punishment (Skinner 1951). Operant H-reflex conditioning allows the patient to self-modulate his/her own monosynaptic spinal reflex activity elicited via constant current electrical stimulation of a peripheral nerve (i.e. H-reflex) (Thompson et al. 2009). The typical procedure involves visual feedback of H-reflex amplitude while the participant operantly learns how to control the feedback signal over many training sessions. Initial work in animal models shows that operant H-reflex conditioning can modulate descending activity of the corticospinal tract and thereby induce spinal cord plasticity that restores altered motor function (Wolpaw et al. 1983; Wolpaw and Lee 1987; Chen and Wolpaw 1995). At present, operant H-reflex conditioning has shown that individuals with spinal cord injury can learn to self-modulate soleus reflex excitability, inducing plasticity at the spinal level (Manella et al. 2013; Thompson et al. 2013; Thompson and Wolpaw 2021). Further, the ability to operantly condition soleus H-reflex correlates with improvements in walking speed, locomotor symmetry, locomotor EMG activity, and H-reflex modulation during locomotion. For instance, Thompson and colleagues (2013) found that some people with SCI can operantly down-condition the soleus H-reflex to a degree comparable to neurologically intact individuals. Manella et al. (2013) found people with motor-incomplete SCI can modulate reflex excitability through operant conditioning in both directions (up/down) on agonist/antagonist pair (tibialis anterior/soleus). This training resulted in improved walking function on a group level. While operant H-reflex conditioning shows promise for addressing hyperreflexia, it is unclear whether quadriceps H-reflex can be successfully trained, and further, whether those post-strokes can down-condition reflex excitability given potential damage to the corticospinal tract (Chen et al. 2002).

In this study, our goal was to examine the feasibility of operant down-conditioning of the RF H-reflex in a pilot cohort of both healthy and post-stroke individuals. We adapted an operant H-reflex conditioning protocol from previous work which focused on regulating soleus H-reflex and targeted the femoral nerve instead of the tibial nerve to innervate the quadriceps. We presented visual feedback of the electrically evoked RF H-reflex, which appeared as a bar graph. Participants were asked to reduce the bar height below a given threshold and were provided a running score of performance. The experiment consisted of 6 baseline sessions without feedback and 24 feedback training sessions, 2-3 sessions per week over a 3-month period. We expected down-regulation of RF H-reflex would be feasible in both healthy and post-stroke individuals. This study represents a novel paradigm in quadriceps spasticity management. This intervention could lead to a targeted, non-pharmacological and non-invasive treatment for hyperreflexia.

## Methods

### Subjects

A total of 7 participants (5 unimpaired [H1-H5], 2 post-stroke [S1-S2]) were recruited for this study. The participants were four men and three women aged 20-68, where two of the individuals had history of stroke (**Table 1**). Each participant was able to stand for 10-minute intervals unassisted, walk on a treadmill for 10-minutes, and provide informed consent. Exclusion criteria included: lower limb musculoskeletal injury, functionally relevant weight-bearing restrictions, cognitive impairment, vision impairment and must not have taken antispasmodic medication one day prior to the session. All seven people participated in down-conditioning. The study was approved by the University of Texas Institutional Review Board, and all subjects gave informed consent prior to participation. To prevent diurnal variation in H-reflex magnitude (Carp et al. 2006), all training was performed at the same time of the day for each individual. Both post-stroke individuals had right hemiparesis and the experiment was conducted on the impaired side.

**Table 1.**
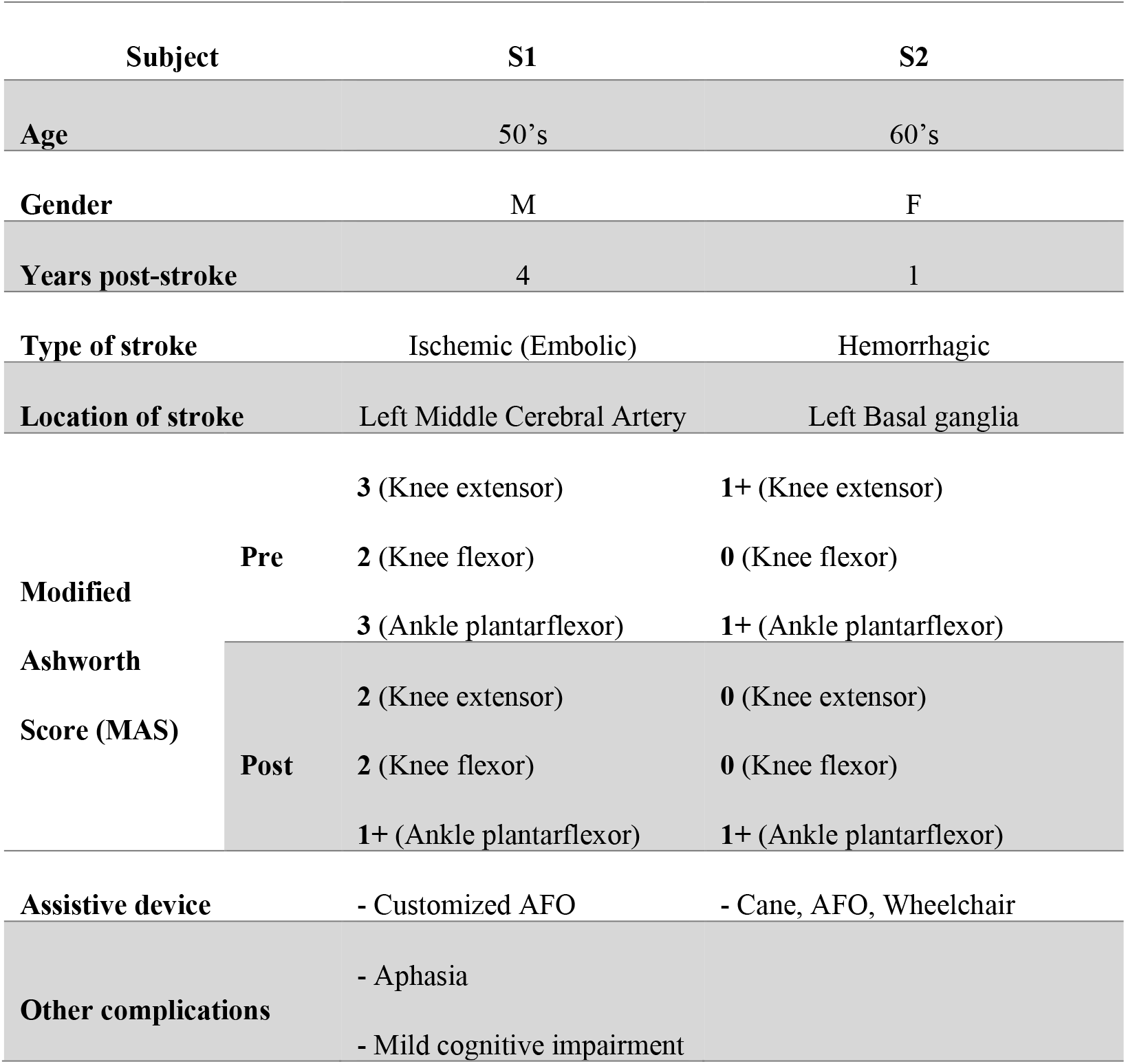
Clinical information of post-stroke participants. Years post-stroke is the duration from the date of stroke to the beginning of the training. Modified Ashworth Score (Bohannon and Smith 1987) was measured by the clinician before and after training. Both participants received therapy outside their experimental participation. MAS Key: 0=no increase in tone, 1=slight increase in tone (catch/release at end ROM), 1+= slight increase in tone, (catch/release at ½ ROM with resistance to end ROM, 2=more marked increase in tone through ROM, passive movement easy, 3=considerable increase in tone, passive movement difficult, 4=affected part in rigid flexion or extension.

### Data collection

The experimental setup consisted of 8-channel surface electromyography (EMG) sensors (AMT-8, Bortec, Calgary, AL), bipolar electrodes (Ag-AgCl, Noraxon, Scottsdale, AZ) a constant current electrical stimulator (Digitimer DS8R, Hertfordshire, UK), a data acquisition board (NI-PCIe 6321, Austin, TX), a desktop computer, and EPOCS software (Devetzoglou Toliou 2020) (**Fig 1**). Additionally, during assessment sessions, stroke subjects walked on an instrumented split-belt force treadmill (Bertec, Columbus, OH), during which the ground reaction forces (GRF) were recorded at 1kHz, and lower limb kinematic data were recorded via inertial motion capture (IMU, Xsens, Enschede, Netherlands) at 60Hz.

**Fig 1.**
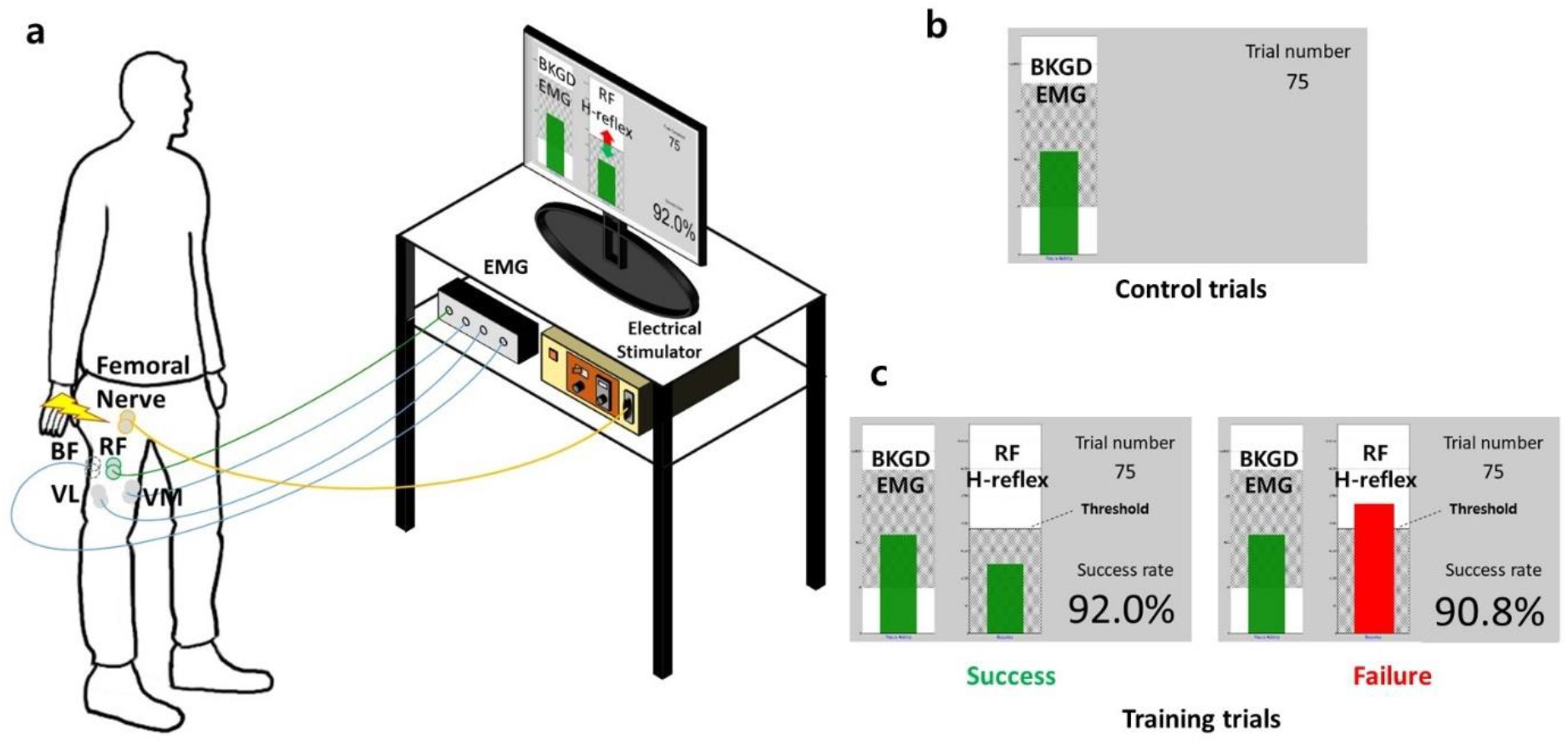
Experimental setup and visual feedback of RF operant H-reflex conditioning. **(a)** Participant with electrodes on the right leg is in the upright standing posture, facing visual feedback from a computer monitor. **(b)** During the control session, only the background EMG level of the RF was provided on the left bar graph. If the background EMG level was within the shaded region and medial hamstring (MH) activity was within a predetermined window, the bar remained green and the electrical stimulation was delivered. Otherwise, the bar turned red with no stimulation or trial progression. **(c)** During the training session, along with control session’s visual feedback, RF H-reflex size and cumulative success rate were provided. The bar turned green if the RF H-reflex was below the threshold and turned red if larger than the threshold.

### Experimental protocol

The experimental protocol consisted of the preparation stage, femoral nerve navigation, recruitment stage, 6 baseline sessions, 24 training sessions, and an additional 2 assessment sessions (pre/post) only for the post-stroke individuals. The experimental sessions occurred 3 times per week over a 12-week period for each individual, where each session lasted 1-1.5 hrs. Each session consisted of preparation, navigation, recruitment, and stimulation stages. The preparation stage started with drawing a grid on a surface of the femoral triangle on the leg. The femoral nerve was identified using an anatomical landmark-based method known as the “four-three finger technique” (Vloka et al. 2018). Surface EMG electrodes were applied over the muscle belly of the rectus femoris (RF), medial hamstring (MH), vastus medialis (VM), and vastus lateralis (VL) based on commonly used anatomical landmarks (Kasman and Wolf 2002; Bordlee and Wong 2015). RF was the primary focus for H-reflex monitoring, MH was monitored for antagonist activity, and VM and VL were monitored to validate the training specificity of the RF H-reflex conditioning. EMG activity was amplified, bandpass filtered (10-1000Hz), and sampled at 1kHz.

During the femoral nerve navigation, the optimal stimulation location of the femoral nerve was established via a monopolar probe electrode to each point on the grid drawn on the femoral triangle (Knikou 2006). Electrical stimulation was delivered as a square stimulus pulse, 1 ms in duration, intensity ranging between 10-20 mA or above depending on the individual, to elicit a reflex response with the subject in quiet standing. We monitored the time-course of the reflex response. The H-reflex was determined as the peak response following the initial stimulus artifact (<3 ms), and M-wave (∼10-25 ms), typically within 30-40 ms post-stimulus. We used the following criteria to identify the optimal H-reflex response: 1) a clear sinusoidal shape, 2) a clear distinction from the M-wave or sinusoidal signal lies on the downward slope of the M-wave in case of contamination, and 3) largest signal magnitude of all the candidate spots. After the optimal stimulation spot was located, the monopolar probe was replaced with a disposable Ag/AgCl snap electrode (circular, 1cm diameter) and the anode electrode (square 5×5cm) was placed over the surface of gluteus maximus (GMax) on the ipsilateral side. To avoid session-to-session variability in the electrodes’ locations, the positions of all electrodes were marked using semi-permanent surgical skin marker. After electrodes were placed, RF maximum voluntary contraction (MVC) was measured. Participants were asked to sit on a chair with their ankle fastened with a belt to the leg of the chair and knee flexed to 90°. The investigator asked the participants to extend their knee with maximum effort for 5-7 seconds. The participant repeated this three times with 15 seconds intervals in between. The representative MVC was determined as mean of three MVC trials and was used to determine the inactive window (15% MVC) for the RF background EMG.

During the recruitment stage, the RF H-reflex recruitment curve was achieved while the subject was in a quiet standing posture. Electrical stimulation with a fixed interval (7 sec) was delivered, only if the background EMG level of these two muscles were below a predetermined threshold of 15% MVC (Pierrot-Deseilligny and Burke 2005). Stimulation intensity was gradually increased by 2mA increments from 10mA to the intensity when the RF M-wave amplitude was the same for three successive trials (Palmieri et al. 2004). For each stimulus intensity, two stimulations were recorded and the mean of two H-reflexes was taken as representative reflex size. The corresponding intensity (I_max_) that elicited maximal representative H-reflex (H_max_) was used in the following trials. Compared to a distal muscle, such as the soleus muscle, the RF H-reflex is often affected by overlap of the M-wave. This overlap makes it difficult to isolate the H-reflex. To address this issue, we used the best decaying exponential fit beginning at the peak of the concurrent M-wave and ending at the zero crossing, then subtracted from the measured signal to obtain the H-reflex estimate (Akbas et al. 2020).

During the baseline/training sessions, surface electromyographic (EMG) activity was amplified, bandpass filtered (3-3000Hz), and recorded from four muscles (RF, MH, VM, and VL), while the femoral nerve was stimulated with the pre-determined intensity (I_max_) at a frequency of 0.14Hz. Every 100ms post-stimulation, peak-to-peak magnitude EMG activity was measured, and the result was provided as visual feedback to the participant. As shown in **Fig 1c**, visual feedback consisted of 1) RF and MH background EMG (left bar), 2) RF H-reflex (right bar), 3) trial number, and 4) cumulative success rate. The left bar was green if the background EMG was below activation threshold but was red otherwise. The goal for the participant was to reduce the height of the right bar, representing H-reflex amplitude, below a given threshold (shaded area in **Fig 1c**). If below the performance threshold, the right bar turned green and cumulative success rate was adjusted accordingly. If above the threshold, the bar turned red and success rate fell. Each trial was 7 s duration, with stimulation of 1ms pulse width followed by feedback for the remainder of the trial. The 6 baseline sessions each consisted of 3 runs of 75 control trials (225 total), while each of 24 training sessions consisted of 20 control trials and 3 runs of 75 training trials (245 total) in standing posture. Subjects were exposed to H-reflex feedback only during the training trials, and no H-reflex feedback was given during the control trials. The participant’s score for each run was cumulative success rate 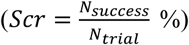 and participants were rewarded in proportion to one’s score (i.e., *Reword*($) = 1.0 + 0.05 * (*Scr* − 50)). If the participant was successful in achieving three consecutive success rates that were equal to or higher than 90% for all three runs, they were provided with an extra dollar as bonus, which made the maximum additional compensation amount $10.00. A rest period was provided between each run to minimize fatigue, which could affect muscular responses to stimuli (Crone et al. 1999; Stutzig and Siebert 2017).

The two individuals with stroke participated in pre/post assessment sessions. Participants performed a 10 m walk test (10MWT) with instructions to walk with maximum effort for 10 m. This process was repeated for 4 times and the participant’s gait kinematics were recorded using inertial motion capture. Next, participants completed the quadriceps pendulum test, which is a non-invasive biomechanical method of evaluating spasticity using gravity to provoke muscle stretch reflexes during passive swinging of the lower limb (Fowler et al. 2007). Inertial measurement units were attached on top of thigh, shank, and foot to calculate the knee joint angle during the process. This test was repeated 3 times with 30 second rest periods in between. The main outcome of this test was quadriceps reflex threshold angle (QRTA), which was the angle difference from the maximum knee extension position to the first swing excursion, the first transition period of knee flexion to knee extension. Third, the participant was asked to walk on an instrumented treadmill while their gait kinematics, ground reaction forces (GRF), and H-reflexes were obtained. During treadmill walking, four gait phases (heel-strike, toe-off, mid-stance, and mid-swing) were detected in real-time using online GRF data. The H-reflex was elicited and measured only when a desired gait phase was detected and occurred at least 7 seconds after the last H-reflex trial. Twenty H-reflexes were recorded for each gait phase. A 5-minute break after each of four runs was provided to minimize the effect of fatigue on H-reflex. The main outcomes consisted of 1) knee flexion range of motion (RoM), 2) peak knee flexion velocity, and 3) H-reflex amplitude during the four different gait phases.

### Outcome Measures

We normalized the H-reflex measurements. For each session, the H-reflex was normalized with the session’s maximum motor response (M-wave, *M*_*max*_), which is the most commonly used method of normalization (Hopkins and Ingersoll 2000). To directly compare between session-to-session performances, each session’s H-reflex trial was averaged and represented as the percentage of the mean of six baseline session’s H-reflex. Main outcome measures of each session were the average of 225 peak-to-peak H-reflex trials normalized to *M*_*max*_ and baseline mean. Control trials, which were conducted without feedback in the beginning of each training session, provide a window into long-term plasticity of H-reflex excitability. For each session, we compared the mean amplitude of 20 control H-reflex trials, which were normalized to the *M*_*max*_ and baseline mean.

### Training specificity of operant H-reflex conditioning

Exploring training specificity of operant H-reflex conditioning can provide evidence that the training can be targeted for a specific muscle, which in this study was the RF. During the experiment, only RF H-reflex visual feedback was provided to the subject while two adjacent quadriceps muscles (VM and VL) were recorded without being visualized by the participant. Since VM and VL share motor innervation with RF through a branch of femoral nerve, we investigated whether the shared nerve root of the quadriceps would project a broader training effect on the quadriceps. H-reflexes of VM and VL were processed in a similar way as RF; each sessions’ H-reflex trials were averaged, normalized by *M*_*max*_, and represented as percentage (%) of 6 baseline sessions’ mean.

### Statistical analysis

We investigated the hypothesis that RF H-reflex can be down-conditioned through operant H-reflex conditioning both in healthy and post-stroke individuals. To validate the effect of training, a paired t-test (*a*<0.05) was used to compare the mean of the 225 normalized H-reflex trials during six baseline sessions to that of the last six training sessions (**Fig 2**). We used a linear mixed model to evaluate any spontaneous change in H-reflex during baseline, where session number was set as a fixed effect and subject number as random effect. Further, we hypothesized RF operant H-reflex conditioning may be targeted to the RF muscle and would not affect other adjacent muscles (VM and VL) using paired t-tests. We compared control H-reflex trials of conditioning sessions to that of baseline sessions to examine the long-term effect of training (Thompson et al. 2009) using a paired t-test.

**Fig 2.**
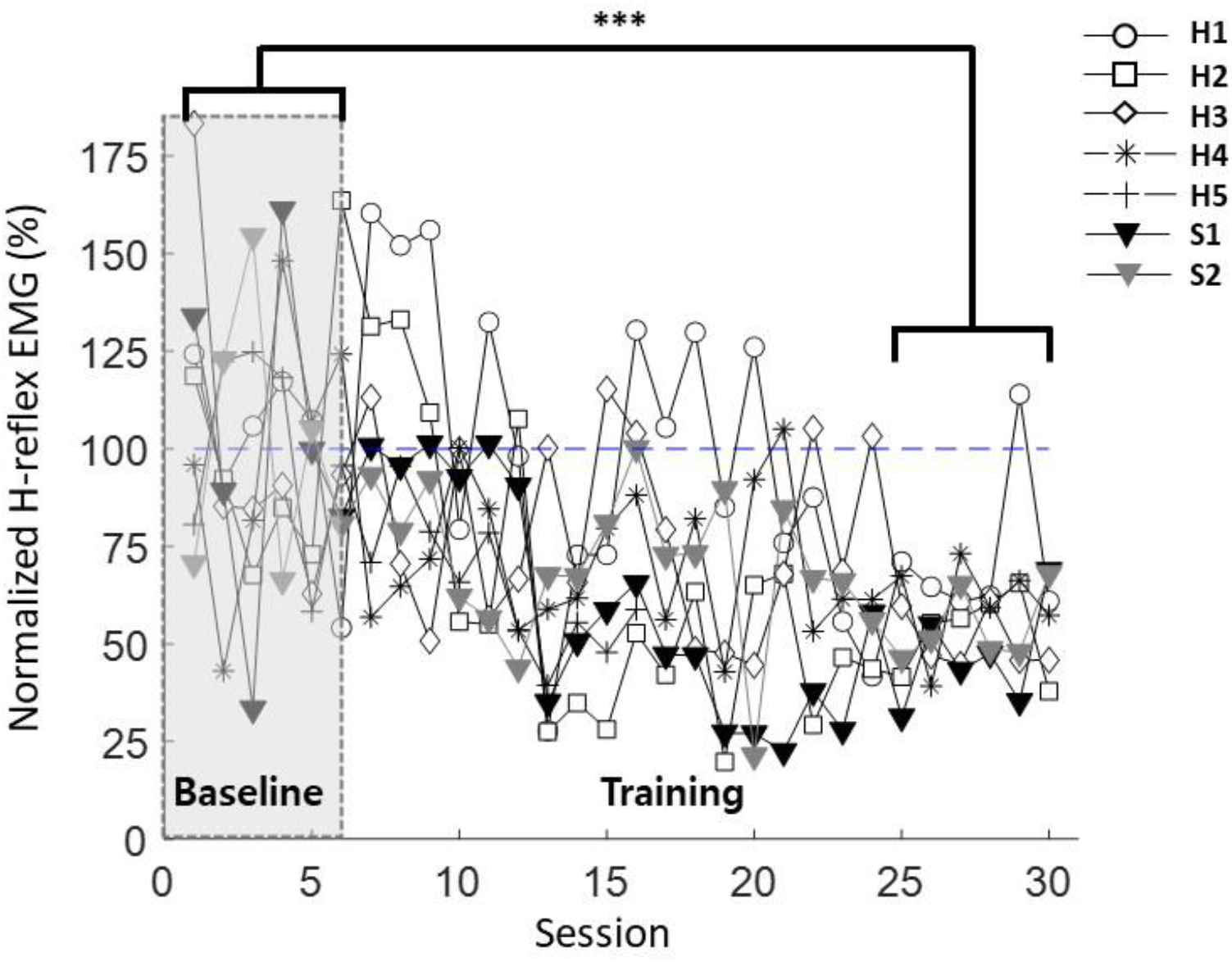
H-reflex magnitude change in RF over training. Shaded area indicates the baseline sessions and blank area indicates training sessions. Each marker represents the individuals’ (H1-H5, S1-S2) RF H-reflex means for each session. (* p<0.05, **p<0.01, ***p<0.001)

For post-stroke individuals, we compared quadriceps reflex threshold angle (QRTA) as a quantitative measure of spasticity, knee joint kinematics, and the H-reflex amplitude during four gait phases (heel-strike, toe-off, mid-stance, and swing) before and after the training using a paired t-test.

## Results

### RF Operant H-reflex conditioning

All participants exhibited the ability to down-regulate RF H-reflex through training. **Table 2** presents the mean response of the healthy participants and individual responses of the two post-stroke participants. Because performance of post-stroke individuals was not significantly different than healthy individuals, the data was pooled for subsequent analyses. There was no spontaneous decrease in H-reflex amplitude during baseline sessions (*F = 0*.*53, p = 0*.*47*), compared to the training sessions (*F = 27*.*18, p < 0*.*0001)*. We observed a decrease in H-reflex amplitude when comparing the last 6 training sessions with the 6 baseline sessions (43.82 ± 4.81% mean±SE, *p < 0*.*0001*). **Fig 2** illustrates the summary statistics of H-reflex amplitude of 7 participants over each session.

**Table 2.**
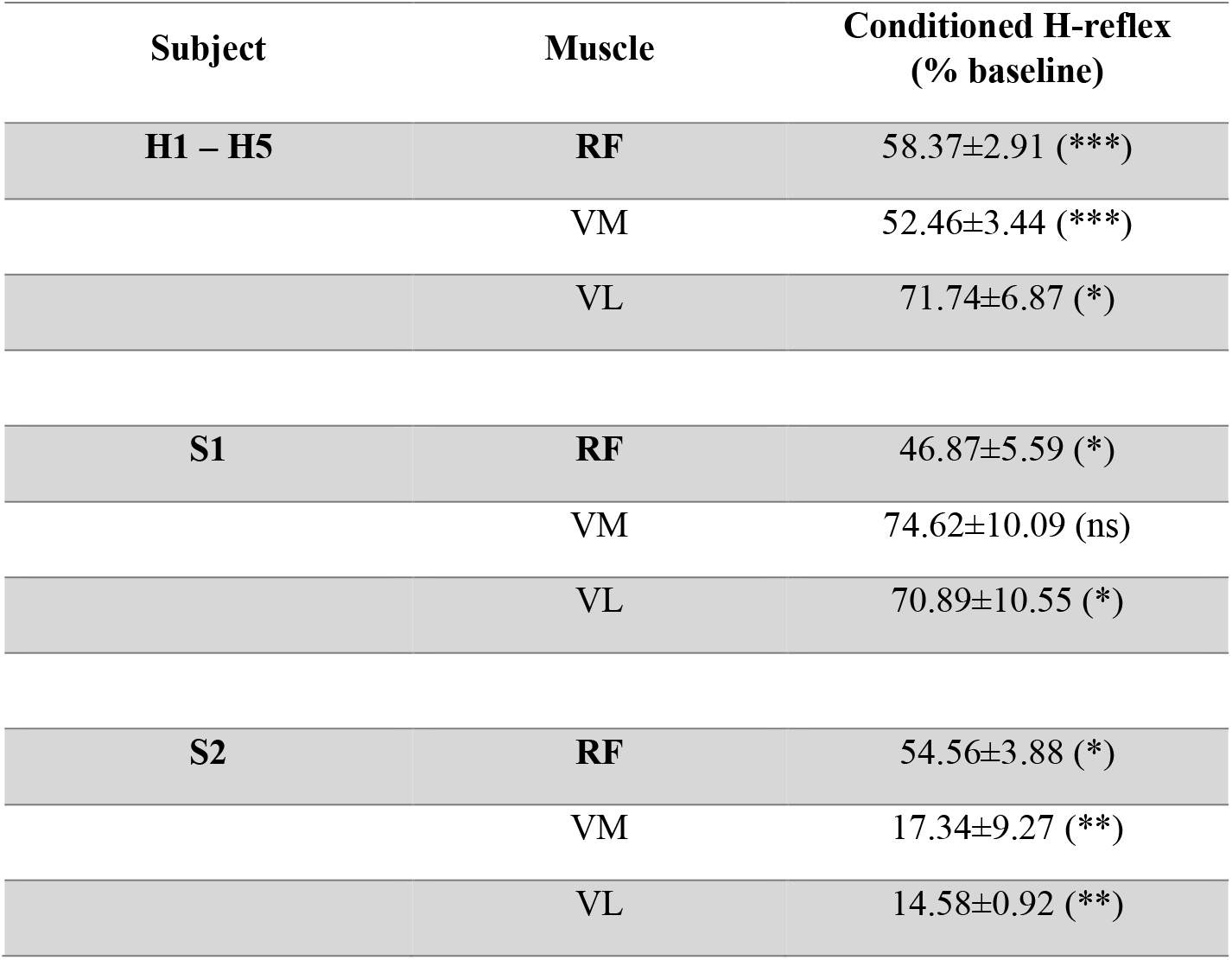
Summary of results based on neurological condition. Values represent mean±SE and are expressed as the percentage of baseline H-reflex mean. The table shows the significant differences from the six baseline sessions’ H-reflex (* p<0.05, **p<0.01, ***p<0.001)

| Subject | Muscle | Conditioned H-reflex<br>(% baseline) |
| --- | --- | --- |
| **H1 – H5** | **RF** | 58.37±2.91 (***) |
|  | VM | 52.46±3.44 (***) |
|  | VL | 71.74±6.87 (*) |
| **S1** | **RF** | 46.87±5.59 (*) |
|  | VM | 74.62±10.09 (ns) |
|  | VL | 70.89±10.55 (*) |
| **S2** | **RF** | 54.56±3.88 (*) |
|  | VM | 17.34±9.27 (**) |
|  | VL | 14.58±0.92 (**) |

### Long-term effect of conditioning on the control H-reflex size

Consolidation of H-reflex down-conditioning following training can be observed in the first 20 control (no feedback) trials of each session. **Fig 3** shows the time-course change of the control RF H-reflex for all participants. For the control trials (i.e., no feedback) of all 7 individuals, the change between baseline and the last 6 training sessions was (20.86 ± 5.77%, mean±SE, *p<0*.*01*).

**Fig 3.**
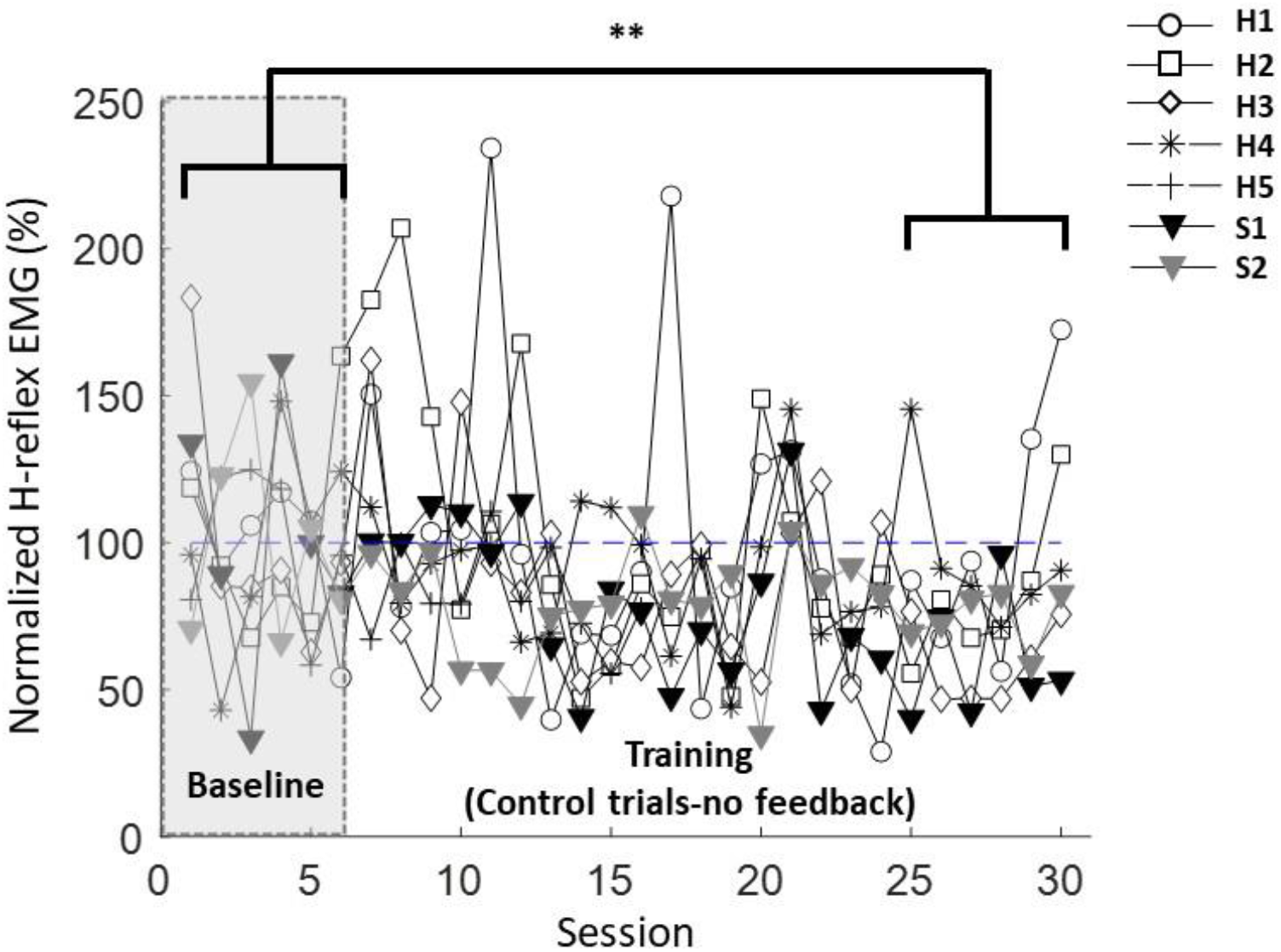
H-reflex magnitude change among control trials shows long-term effect of the training. Data during training phase extracted from 20 control trials prior to introduction of feedback for each session. Each marker represents the individuals’ (H1-H5, S1-S2) control RF H-reflex means for each session. (* p<0.05, **p<0.01, ***p<0.001)

### Training specificity of RF operant H-reflex conditioning: Comparison with VM and VL

The femoral nerve innervates all four quadriceps muscles, but feedback is only provided from one muscle, raising a question of muscle training specificity. Performance of healthy individuals separate from post-stroke individuals can be seen in **Table 2**. For healthy individuals, H-reflex decreased significantly for both VM and VL, whereas mixed result was monitored among post-stroke individuals. We pooled these results in **Fig 4** for VM (49.16± 5.42% (SE) drop, *p<0*.*0001*) and VL (37.24 ± 7.37%(SE) drop, *p<0*.*001*), respectively.

**Fig 4.**
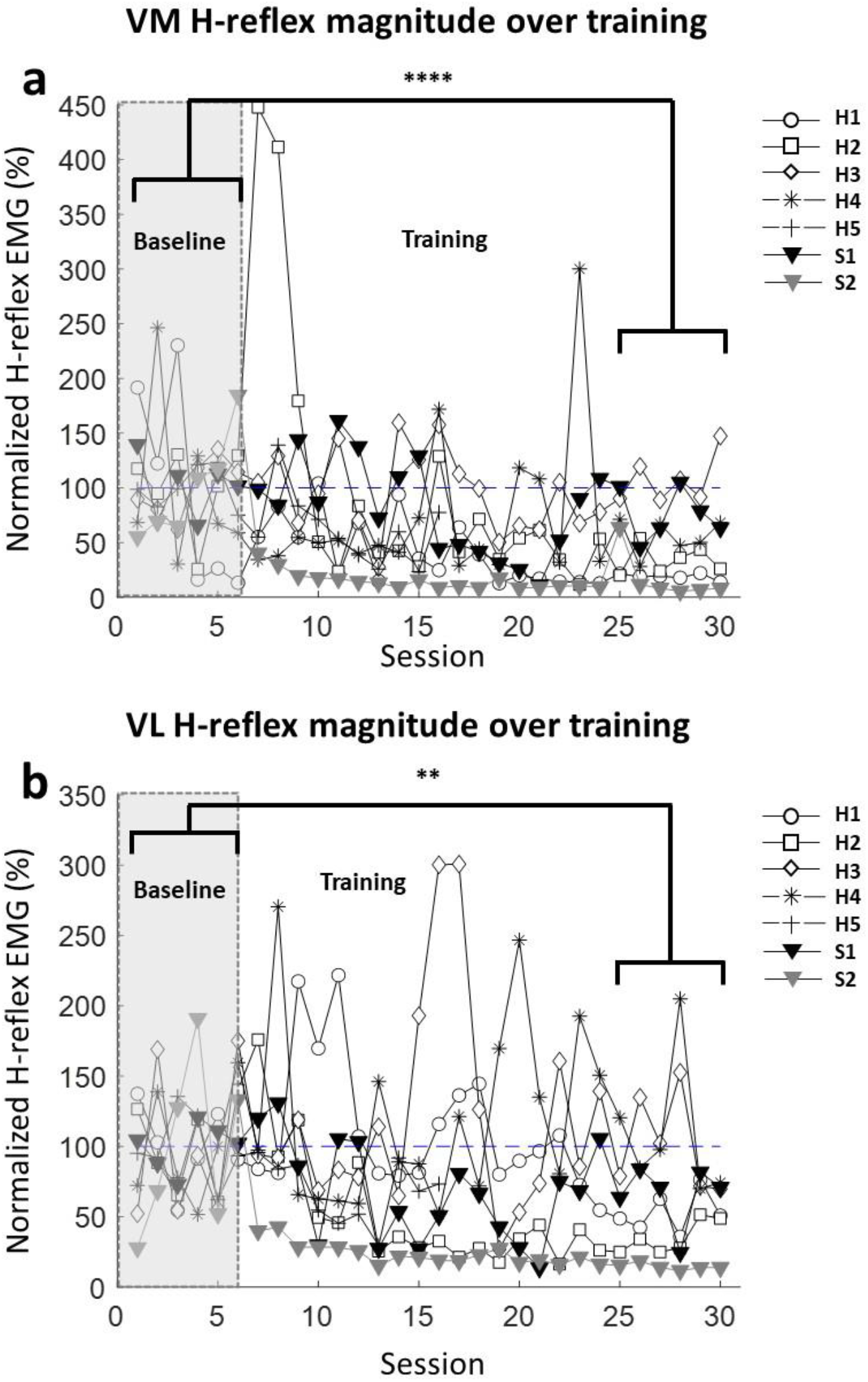
Muscle training specificity of RF Operant H-reflex Conditioning in VM (a) and VL (b) in all participants. Shaded area indicates the baseline sessions and blank area indicates training sessions. Each marker represents the individuals’ (H1-H5, S1-S2) VM and VL H-reflex means for each session. (* p<0.05, **p<0.01, ***p<0.001, ****p<0.0001)

### Behavioral effect of the RF operant H-reflex conditioning in Post-Stroke Individuals

We conducted a pre/post behavioral assessment on the two post-stroke individuals. Knee flexion RoM and peak knee flexion velocity significantly improved both during overground and treadmill walking for S1. However, for S2, the improvement in knee flexion kinematics was limited to RoM during overground walking. **Table 3** illustrates the results. Quadriceps spasticity, quantified as QRTA, improved in S1 by 10.55° (*p=0*.*016*), while S2 had a statistically insignificant change of 3.50° *(p = 0*.*536*). Reflex modulation (**Table 3**) for S1 improved significantly for all four gait phases, where the mean reduction was 19.21%. In S2, H-reflex excitability decreased significantly in three gait phases (heel-strike, toe-off, and mid-stance), with a mean decrease of 23.4%.

**Table 3.**
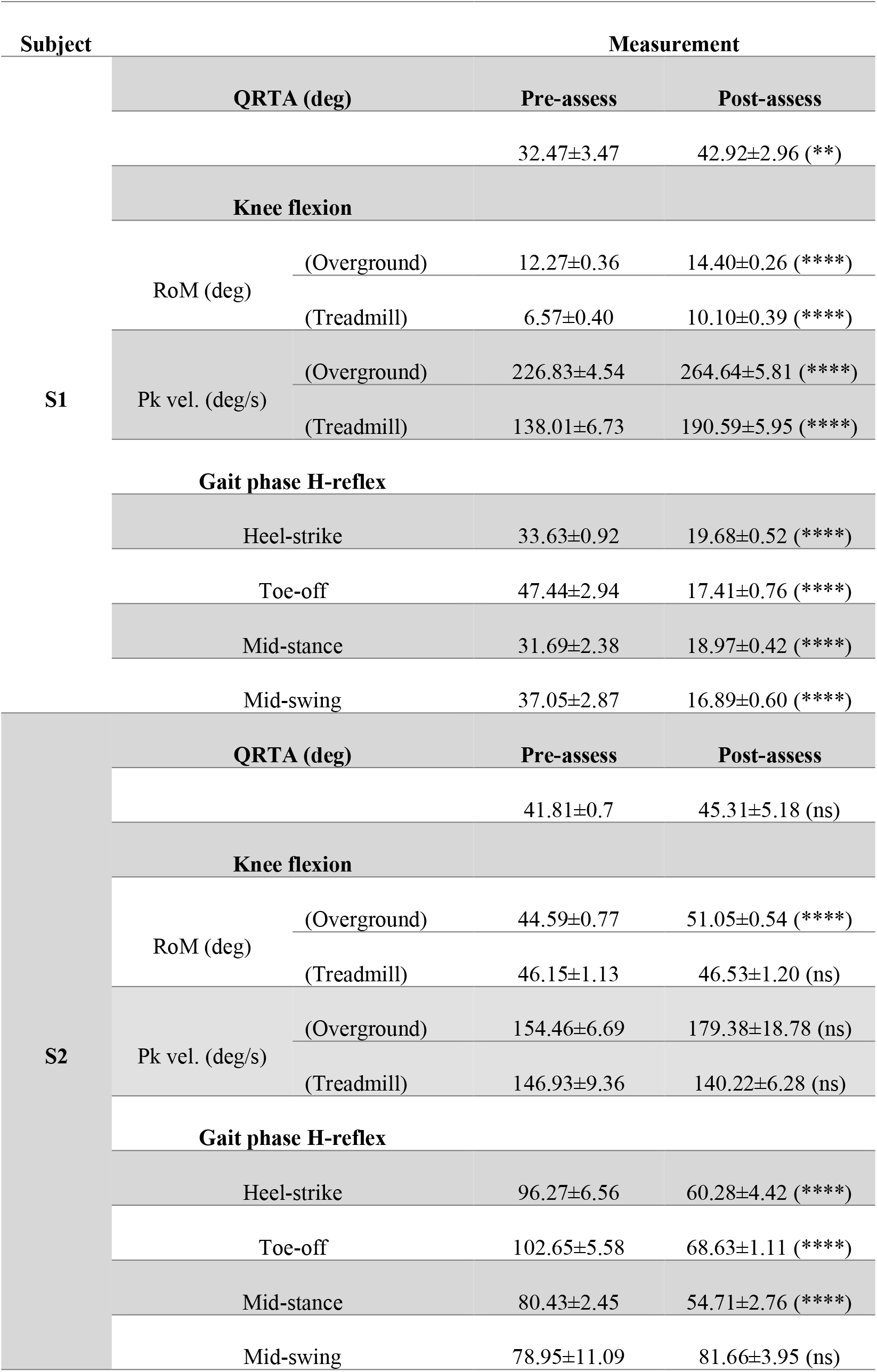
Behavioral effect on gait kinematics. Values represent mean±SE of Quadriceps Reflex Threshold Angle (deg), knee flexion RoM (deg), and peak-velocity (deg/s) during overground (10MWT) walking and treadmill walking. The table shows the significant differences between the pre-assessment and post-assessment session (* p<0.05, **p<0.01, ***p<0.001, ****p<0.0001)

| Subject | Measurement |  |  |  |
| --- | --- | --- | --- | --- |
| S1 | QRTA (deg) |  | Pre-assess | Post-assess |
|  |  |  | 32.47±3.47 | 42.92±2.96 (**) |
|  | Knee flexion |  |  |  |
|  | RoM (deg) | (Overground) | 12.27±0.36 | 14.40±0.26 (****) |
|  |  | (Treadmill) | 6.57±0.40 | 10.10±0.39 (****) |
|  | Pk vel. (deg/s) | (Overground) | 226.83±4.54 | 264.64±5.81 (****) |
|  |  | (Treadmill) | 138.01±6.73 | 190.59±5.95 (****) |
|  | Gait phase H-reflex |  |  |  |
|  | Heel-strike |  | 33.63±0.92 | 19.68±0.52 (****) |
|  | Toe-off |  | 47.44±2.94 | 17.41±0.76 (****) |
|  | Mid-stance |  | 31.69±2.38 | 18.97±0.42 (****) |
|  | Mid-swing |  | 37.05±2.87 | 16.89±0.60 (****) |
| S2 | QRTA (deg) |  | Pre-assess | Post-assess |
|  |  |  | 41.81±0.7 | 45.31±5.18 (ns) |
|  | Knee flexion |  |  |  |
|  | RoM (deg) | (Overground) | 44.59±0.77 | 51.05±0.54 (****) |
|  |  | (Treadmill) | 46.15±1.13 | 46.53±1.20 (ns) |
|  | Pk vel. (deg/s) | (Overground) | 154.46±6.69 | 179.38±18.78 (ns) |
|  |  | (Treadmill) | 146.93±9.36 | 140.22±6.28 (ns) |
|  | Gait phase H-reflex |  |  |  |
|  | Heel-strike |  | 96.27±6.56 | 60.28±4.42 (****) |
|  | Toe-off |  | 102.65±5.58 | 68.63±1.11 (****) |
|  | Mid-stance |  | 80.43±2.45 | 54.71±2.76 (****) |
|  | Mid-swing |  | 78.95±11.09 | 81.66±3.95 (ns) |

## Discussion

The goal of this study was to investigate the feasibility of self-modulation of RF H-reflex excitability. We trained 7 individuals (5 healthy and 2 post-stroke), over 24 sessions. Our results provided the first evidence that RF H-reflex can be down-regulated, and we did not observe any decrement in ability to self-regulate in 2 post-stroke individuals. The down-regulation of RF generalized to other quadriceps muscles (VM and VL). The 2 post-stroke individuals exhibited improvements in reflex modulation generalized over the gait cycle and gait performance. We conclude that RF H-reflex down-conditioning is feasible and its effects should be evaluated in a controlled clinical trial.

Our findings suggest that individuals were successful in down-regulating their RF H-reflex via operant H-reflex conditioning. Comparing the H-reflex amplitude in the six baseline sessions to the last six training sessions resulted in a 42% drop in healthy individuals, 49% drop for the two post-stroke individuals, and 44% drop across all 7 individuals. Previous animal model research indicated that CST transmission was critical for operant H-reflex learning (Chen et al. 2002) suggesting that those with reduced CST integrity resulting from stroke may not be able to learn. Our findings indicate that damaged corticospinal pathways due to stroke do not prevent successful learning. A controlled trial remains to be conducted that examines the effect of damaged pathways on operant learning and performance.

We expected to observe successful down-conditioning of RF since previous work has shown successful soleus H-reflex down-conditioning (Thompson et al. 2009; Manella et al. 2013). However, in neural operant conditioning, even across modalities, approximately 30% of individuals are non-learners (Thompson et al. 2009; Manella et al. 2013; Sitaram et al. 2017). Although only 7 individuals were included in our cohort, there were no non-responders, suggesting the potential for a robust operant conditioning paradigm. Our analysis of 6 baseline sessions showed no evidence of spontaneous decrease in H-reflex amplitude that could be mistaken as down-conditioning. Further, there is no evidence of spontaneous decrease in H-reflex amplitude over sessions in previous work (Thompson et al. 2009). Thus, it is highly unlikely that RF H-reflex spontaneously decreased over time.

Trials conducted without feedback, control trials, provide a window into long-term plasticity. Overall, we observed a decrease in all individuals (21 ± 6(SE) %), where the drop in post-stroke individuals (S1 = 25%, S2 = 40%) was more evident than that of the healthy group (16 %). These results align with earlier findings, where the drop of control H-reflex of the soleus was larger in participants with spinal cord injury (24%) than healthy individuals (16%) (Thompson et al. 2013). This difference is attributed to the hypothesis of negotiated equilibrium (Wolpaw 2018), the concept that neural circuitry already behaving well will not be affected by neuromodulation, but neurologically affected circuitry will.

We found that operant conditioning based on femoral nerve stimulation generalized to other innervated quadriceps muscles. Additional monitoring of adjacent quadriceps muscles (VM and VL) was performed to validate the specificity of RF operant H-reflex conditioning. We found a significant drop in VM and VL in healthy individuals between the first and last 6 sessions. Although not conclusive, these results suggest that operant conditioning based on stimulation of a peripheral nerve that innervates multiple muscles may affect the entire muscle group. Animal studies by Chen et al. (Chen et al. 2011) found that operant conditioning of the soleus affected quadriceps H-reflex excitability, and as such the generalization of operant conditioning to multiple muscles, especially innervated by the same nerve, was expected. The degree of generalization will indicate the specificity of the effect of training and needs to be characterized further.

We found functional changes in gait in the post-stroke individuals following training. These results align with earlier studies that have shown functional improvement following the operant H-reflex conditioning. Chen et al. (2006) have shown rats with incomplete spinal cord injury (SCI) improved gait asymmetry following the training. The studies by Thompson et al. and Manella et al. (Manella et al. 2013; Thompson et al. 2013) have shown gait improvements (i.e., speed, endurance, less clonus, easier stepping etc.) in people with SCI. Most recently, Tahayori and Koceja (2019) have shown 3 post-stroke individuals were able to down-regulate their soleus H-reflex and the changes were correlated with movement improvements. In our study, S1 exhibited improvements in knee ROM and knee flexion velocity in both overground and treadmill walking conditions, but S2 only showed improvements in knee kinematics only in overground walking. Both participants exhibited better improvements during overground walking than treadmill walking. Overground walking was measured during 10m walk test, where the participants were asked to set their speed with the maximum effort. No wiring from EMG sensors or the stimulator was attached, and a belt type harness was worn. In contrast, during treadmill walking, participants had to walk in a fixed and self-selected speed that was predetermined, the wiring from EMG sensors and stimulator was present, a full-body harness had to be worn, and no gait support devices (i.e., cane, rolling walker) were allowed. Thus, it is possible that the differences between overground and treadmill walking could be due to the experimental setup. Clinical and laboratory measurements of hyperreflexia showed improvements in both post-stroke individuals. S1 improved in QRTA, while we did not observe a change in QRTA of S2. Further, H-reflex amplitude during different gait cycle phases also decreased, suggesting generalization of training during standing to walking (Thompson and Wolpaw 2021). While no conclusions can be drawn across post-stroke individuals, these results suggest the efficacy of conducting a RF H-reflex down-conditioning clinical trial for individuals post-stroke with SKG and quadriceps hyperreflexia.

This study investigates the feasibility of RF H-reflex down-conditioning based on 7 individuals, 2 of them post-stroke. One of the main limitations of this study is the small sample size (7). Despite the small sample, the effect size (3.85) and statistical power (98.7%) were substantial and suggest that running more individuals would not change our conclusions. Due to training only 2 post-stroke individuals, we cannot generalize the feasibility in operant conditioning of RF H-reflex conditioning to a larger population. It is interesting that S2 demonstrated improvement in H-reflex during gait phases, except for mid-swing, yet did not show as great of a functional improvement as S1. While the S2 mechanism (cortico-basal ganglia-thalamo-cortical loop pathway) underlying this change was different than, S1 (corticospinal pathway), it is notable that both participants successfully down-regulated RF H-reflex. Further investigations should be dedicated to understanding the role between H-reflex excitability and clinical outcomes.

## Conclusions

Our observations suggest that RF H-reflex down-conditioning is feasible and generalizes to other quadriceps muscles. The successful training of 2 post-stroke individuals provides promise that one’s ability to regulate H-reflex may extend to this neurologically impaired population, but a larger sample size is needed to confirm feasibility, long-term plasticity and clinical effects. These initial results open the door for alternative treatments of spasticity and may lead towards an effective intervention in post-stroke Stiff-Knee gait and other spasticity-related disorders.

## Data Availability

All data produced in the present study are available upon reasonable request to the authors

## Acknowledgements

We would like to thank the participants for their efforts. We would also like to thank Dr. Amir Eftekhar and Dr. Aiko Thompson for developing Evoked Potential Operant Conditioning System (EPOCS) software and providing helpful guidance.

